# Characterizing Post-stroke Gait Propulsion Beyond Walking Speed: A Clinically Feasible Approach Using the Functional Gait Assessment

**DOI:** 10.1101/2025.11.03.25339246

**Authors:** Jeffrey Paskewitz, Jie Fei, Ruoxi Wang, Louis N. Awad

**Author notes:** These authors contributed equally to this work (Jeffrey Paskewitz, Jie Fei).

## Abstract

Post-stroke gait dysfunction is biomechanically heterogeneous, yet biomechanically-informed classifications of functional walking remain underdeveloped. In particular, there is a lack of clinically accessible methods for classifying gait deficits that account for propulsion impairments—a historically laboratory-dependent gait parameter requiring measurement with force plate systems. This study examined whether propulsion impairment can be classified by combining a global measure of walking function (i.e., the 10-meter walk test speed) with specific measures of dynamic walking ability derived from individual items of the Functional Gait Assessment (FGA). Forty participants >6 months poststroke completed biomechanical evaluations quantifying propulsion during walking and clinical assessments including the FGA. Multivariable stepwise regression identified the FGA items most strongly associated with paretic propulsion. Models augmented with these FGA items explained 10-14% greater variance in propulsion peak and 2-5% greater variance in propulsion impulse compared with models using walking speed alone. Incorporating FGA items also yielded the highest overall accuracy (72.5%) and per-class performance in propulsion severity classification. These findings establish the co-assessment of walking speed and targeted FGA items as a clinically-feasible approach to biomechanically-informed classification of post-stroke gait dysfunction.

## 1. Introduction

Stroke affects more than 795,000 individuals annually in the United States [1] and remains one of the leading causes of long-term disability worldwide. The direct and indirect costs of stroke amount to an estimated $56 billion per year in the United States alone [1]. Mobility is reduced in more than half of survivors aged 65 and older [2]. Given its strong association with independence and quality of life, recovery of walking function is a central goal of rehabilitation [3-6]. However, although rehabilitation interventions can improve walking after stroke, outcomes remain highly variable [7-13].

A major contributor to this variability is the biomechanical heterogeneity of post-stroke gait impairment. The most common classification approach used to examine the effects of rehabilitation is based on baseline deficits in walking speed, where cutoffs of 0.80 m/s [14] and 0.93 m/s [15] have been proposed to differentiate limited and unlimited community ambulators. Although this approach is valuable, it overlooks the distinct biomechanical strategies individuals use to achieve a given speed [14]. Indeed, any two individuals post-stroke who walk at similar speeds may rely on profoundly different locomotor patterns to compensate for distinct biomechanical deficits. Prior work suggests that predictions of an intervention’s therapeutic effects improve substantially when both walking speed and gait biomechanics are considered [16]. This underscores the need for biomechanically informed classification of post-stroke gait dysfunction, with emphasis on approaches that can be implemented clinically.

Propulsion is a key biomechanical subtask that enables stable and efficient forward progression during gait [17-21]. Propulsion arises from the interaction of kinematic and kinetic factors that are difficult to measure in clinical settings, historically requiring laboratory-based motion capture and force plate systems. Although experienced clinicians can grossly estimate propulsion deficits based on walking speed and observable (i.e., kinematic and spatiotemporal) gait deviations [22, 23], observational gait analysis has considerable limitations [24-26]. While very large propulsion deficits may be visually apparent, clinically-accessible methods for classifying walking deficits based on subtle yet meaningful deficits in gait propulsion are lacking [16, 27, 28]. Such methods are essential for guiding intervention selection and detecting treatment-related changes [16, 28].

The Functional Gait Assessment (FGA) is a reliable and valid measure of dynamic walking function that evaluates ten ambulatory tasks, each scored from 0 (severe impairment) to 3 (no impairment) [3, 22-33]. Like walking speed, the FGA is feasible to administer in routine clinical practice and provides insight into gait quality, suggesting potential value in characterizing biomechanical impairments [30]. The purpose of this study was to examine the extent to which co-assessment of walking speed and the FGA can characterize propulsion impairment after stroke. We hypothesized that walking speed and specific FGA items together would improve the classification of propulsion impairment beyond walking speed alone, supporting the development of a clinically feasible, biomechanically-informed framework for walking assessment.

## 2. Materials and Methods

### 2.1. Recruitment

Forty individuals were recruited from a registry of individuals who had previously sustained a stroke and expressed interest in participating in research studies. The inclusion criteria were: age 18 years or older, prior history of stroke, medically stable, ability to walk without another individual supporting the person’s body weight for at least two minutes (assistive devices such as a cane were allowed), ability to communicate with investigators and follow instructions, and medical clearance provided by a physician. Exclusion criteria were: inability to communicate, a score >1 on question 1b and >0 on question 1c of the NIH Stroke Scale, pain that impaired walking ability, presence of neglect and hemianopia, unexplained dizziness in the last six months, serious comorbidities that may interfere with participation (musculoskeletal, cardiovascular, pulmonary, and neurological), and history of more than two falls in the previous month. All study procedures were approved by the Boston University Institutional Review Board. Written informed consent and physician medical clearance were obtained from all participants.

### 2.2. Data Collection

During the study visit, each participant completed the FGA, three trials of the time-scored 10-meter walk test (10MWT) at a comfortable speed, and one trial of the 6-minute walk test (6MWT). The FGA and Fugl-Meyer Assessment for the Lower Extremity (FMA-LE) were administered and scored by licensed physical therapists. For the 10MWT, participants walked along a 10-meter straight section of a 26.6-meter oval indoor track. For the 6MWT, participants walked around the entire track while ground reaction forces (GRF) were captured each time they passed the straight section containing six floor-embedded force plates (Bertec Corp., Columbus, OH, USA). GRF data for both legs were collected at a sampling rate of 2000 Hz. The use of assistive devices was allowed during all assessments, if needed.

### 2.3. Data Processing

Raw GRF data collected during the 6MWT were exported from Qualisys Track Manager Version 2024.3 (Qualisys AB, Göteborg, Sweden) and processed using MATLAB Version R2024a (MathWorks Inc., Natick, MA). A second-order low-pass Butterworth filter with a cutoff frequency of 10 Hz was applied to filter the raw GRF data. Gait events of heel strike and toe-off were identified based on a force threshold of 5% body weight (BW). GRF data were then segmented from heel strike to toe-off to represent the stance phase of walking [34]. Any segmented GRF data identified as abnormal due to cross-over strides on the force plates were excluded. Anteriorly directed GRF for both legs was extracted to compute three gait propulsion point metrics, including the paretic propulsion peak, paretic propulsion impulse, and propulsion impulse symmetry [35]. Paretic propulsion peak was calculated as the maximum anteriorly directed GRF normalized by body weight, reported in the unit of %BW, while the paretic propulsion impulse was calculated as the integral of the anteriorly directed GRF normalized by body weight [34]. Propulsion impulse symmetry was determined using equation (1) below:

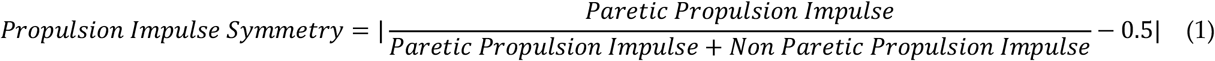

Data points were identified as outliers and excluded if they exceeded three standard deviations from the mean. The remaining point metrics data were then averaged for analysis. Higher paretic propulsion peak and impulse, and lower propulsion impulse symmetry indicate less severe gait impairment [14, 16]. The CWS was calculated as the average gait speed across three 10MWT trials. Gait speed for each trial was calculated by dividing the distance of steady state walking (6 meters, excluding the acceleration and deceleration phases) by the time taken to complete the distance.

### 2.4. Data Analysis

All data analyses and figure generation were performed in MATLAB Version R2024a (MathWorks Inc., Natick, MA). Pearson correlations were calculated to examine associations between propulsion point metrics and both the FGA total score (FGA Total) and CWS. Univariable linear regression models were first constructed with CWS as the sole predictor for each of the three propulsion metrics: paretic propulsion peak, paretic propulsion impulse, and propulsion impulse symmetry. To evaluate whether the inclusion of FGA statistically improved model performance, two multivariable regression models were developed to estimate the same propulsion point metrics using: (1) CWS and the FGA Total, and (2) CWS in combination with individual FGA items selected via stepwise regression based on the Bayesian Information Criterion (BIC) [36].

Two pairs of nested models were compared using F-tests: the univariable model (CWS-only) versus the multivariable model 1 (CWS + FGA Total), and the univariable model versus the multivariable model 2 (CWS + stepwise-selected FGA items). Variance inflation factors (VIFs) were calculated to assess multicollinearity among predictors, with variables exceeding a VIF of 5 removed. Model performance was evaluated using R-squared (R^2^), adjusted R^2^, Root-Mean-Square Error (RMSE), and model-level *p*-values. Regression coefficients and corresponding *p*-values were reported for all retained predictors. F-statistics, degrees of freedom, and *p*-values from the F-tests were reported for nested model comparisons. Model generalizability was assessed using 10-fold cross-validation, with cross-validated R^2^ and RMSE reported.

To further examine whether FGA-augmented models improved the classification of propulsion severity beyond CWS alone, participants were divided into three hemiparetic severity groups (severe, moderate, and mild) based on tertiles of force-plate-derived paretic propulsion peak. Group differences in paretic propulsion peak and impulse were tested using one-way ANOVA, followed by Bonferroni-corrected post hoc comparisons. Predicted paretic propulsion peak values from each of the three regression models were then used to classify participants into severity groups using the same tertile cutoffs. These predicted classifications were compared with force-plate-derived ground-truth classifications. Model classification performance was summarized using overall accuracy, confusion matrices, and class-specific sensitivity, specificity, and precision.

## 3. Results

Eleven female and twenty-nine male participants completed the study. In two participants, due to time constraints, the FGA was performed during a second study visit completed within 7 days of the first study visit. All other participants completed the study according to the described methods. Participant demographics are summarized in Table 1. The mean (± SD) age was 61.52 ± 9.95 years, with a BMI of 28.77 ± 5.79 kg/m^2^, and stroke chronicity of 7.97 ± 4.18 years. Twenty-two participants presented with right-sided paresis, and eighteen with left-sided paresis. The FMA-LE score was available for 36 participants, with a mean of 18.47 ± 5.02; the mean CWS was 0.85 ± 0.29 m/s, and the mean FGA Total was 16.62 ± 6.09. During FGA, eight participants used an AFO or AD, twenty-nine ambulated without support, and usage data were unavailable for three participants.

**Table 1.** Participant demographics.

| Characteristic |  |
| --- | --- |
| Age (sd), y | 61.52 (9.95) |
| BMI (sd), kg/m<sup>2</sup> | 28.77 (5.79) |
| Chronicity (sd), y | 7.97 (4.18) |
| Sex (M/F) | 29/11 |
| Paretic leg (L/R) | 22/18 |
| FMA-LE (sd) * | 18.47 (5.02) |
| CWS (sd), m/s | 0.85 (0.29) |
| FGA Total (sd) | 16.62 (6.09) |
| AFO or AD using during FGA, (yes/no/missing) ** | 8/29/3 |

\* FMA-LE data were missing for 4 participants
\*\* AFO and AD usage data were missing for 3 participants
**Abbreviations:** sd = standard deviation; y = year; BMI = Body Mass Index; FMA-LE = Fugl-Meyer Assessment for Lower Extremity; CWS = Comfortable Walking Speed; FGA = Functional Gait Assessment; AFO = Ankle-Foot Orthosis; AD = Assistive Device

The distribution of propulsion point metrics is illustrated in Figure 1a. The paretic prolusion peak averages 9.37 ± 5.81 %BW, ranging from -0.72 to 21.06 %BW. The paretic propulsion impulse is 2.46 ± 1.42 %BW, with values ranging from 0.01 to 5.19 %BW. The propulsion impulse symmetry shows a mean of 0.17 ± 0.15, with values ranging from 0.00 to 0.50. A stacked bar plot showing the frequency of participant scores (0-3) across 10 items of the FGA is shown in Supplementary Figure A1. Figure 1b illustrates moderate positive correlations between CWS and both paretic propulsion peak (r = 0.68, p < 0.001) and paretic propulsion impulse (r = 0.68, p < 0.001) and propulsion impulse symmetry (r = -0.50, p < 0.001) [36]. Figure 1c shows similar patterns for the FGA Total. A strong positive correlation is observed between FGA Total and paretic propulsion peak (r = 0.73, p < 0.001), and moderate correlations are observed between FGA Total and both paretic propulsion impulse (r = 0.65, p < 0.001) and propulsion impulse symmetry (r = -0.48, p < 0.01).

**Figure 1.**
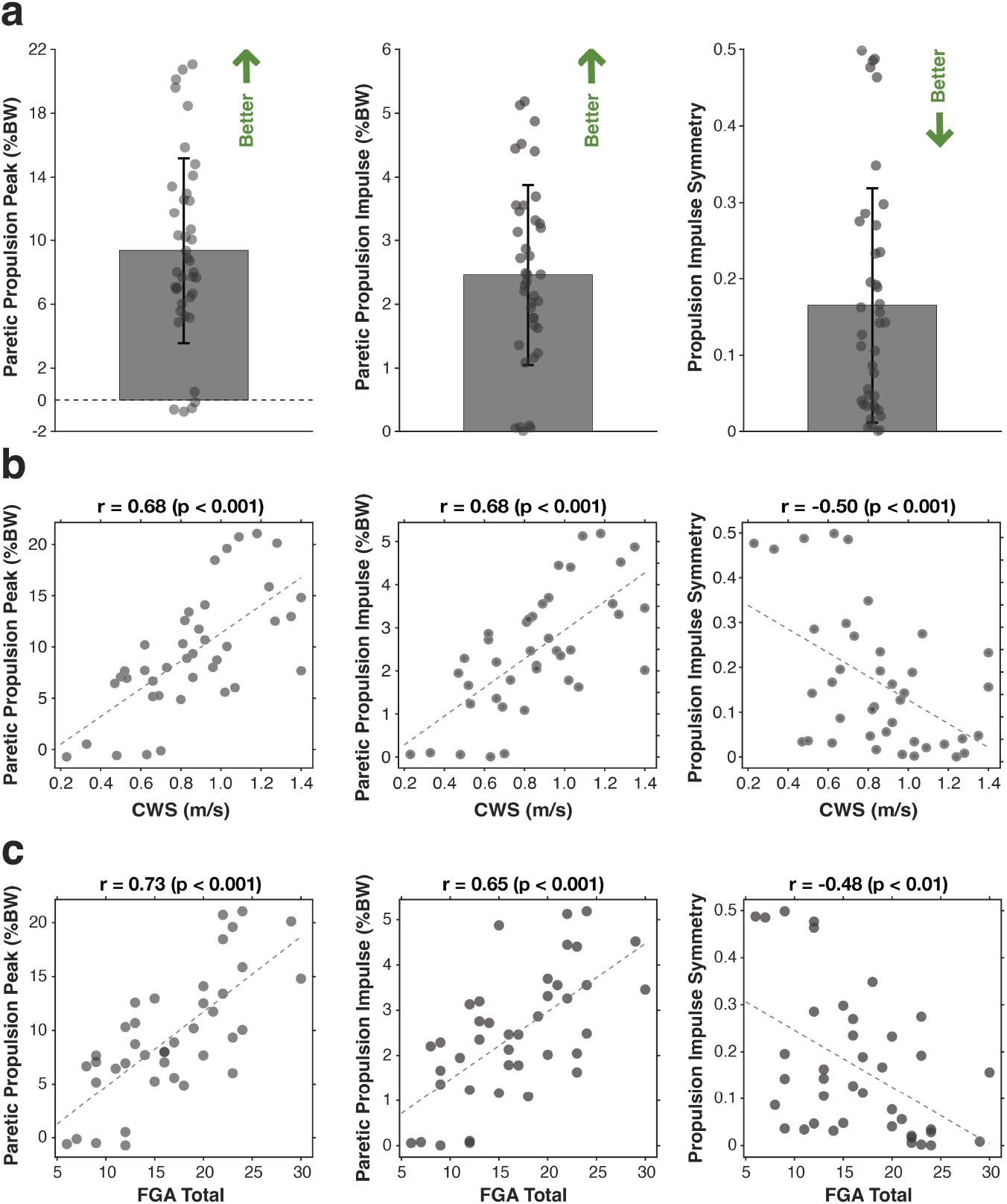
Distribution of propulsion point metrics and their Pearson’s correlation analysis with CWS and FGA Total : **(a)** Distribution of paretic propulsion peak, paretic propulsion impulse, and propulsion impulse symmetry among participants. Green arrows point toward better propulsion performance; **(b)** Correlation between each propulsion point metric and CWS; **(c)** Correlation between each propulsion point metric and FGA Total.

Regression analysis of the paretic propulsion peak is summarized in Table 2. Univariable regression using CWS as the sole predictor explains 45% of the variance in paretic propulsion peak (Adj R² = 0.45, RMSE = 4.32 %BW, p < 0.001), demonstrating a significant positive relationship between CWS and paretic propulsion peak (β = 13.55, p < 0.001). Adding the FGA Total in multivariable model 1 improves the explained variance to 56% (Adj R² = 0.56, RMSE = 3.87 %BW, p < 0.001), with the FGA Total emerging as a significant predictor (β = 0.48, p < 0.01), and significantly improving model performance compared to the CWS-only model (F(1, 37) = 10.38, p < 0.01). Multivariable model 2 using stepwise-selected FGA items (FGA 7 and FGA 10) yields the best model performance, explaining 60% of the variance (Adj R² = 0.60, RMSE = 3.68 %BW, p < 0.001). This model also significantly improves model performance compared to the CWS-only model (F(2, 36) = 8.26, p < 0.01). Both selected FGA items are statistically significant (p < 0.05), along with CWS (β= 8.31, p < 0.01). Cross-validation confirms the improved performance of this model (CV R² = 0.57, CV RMSE = 3.77 %BW).

**Table 2.**
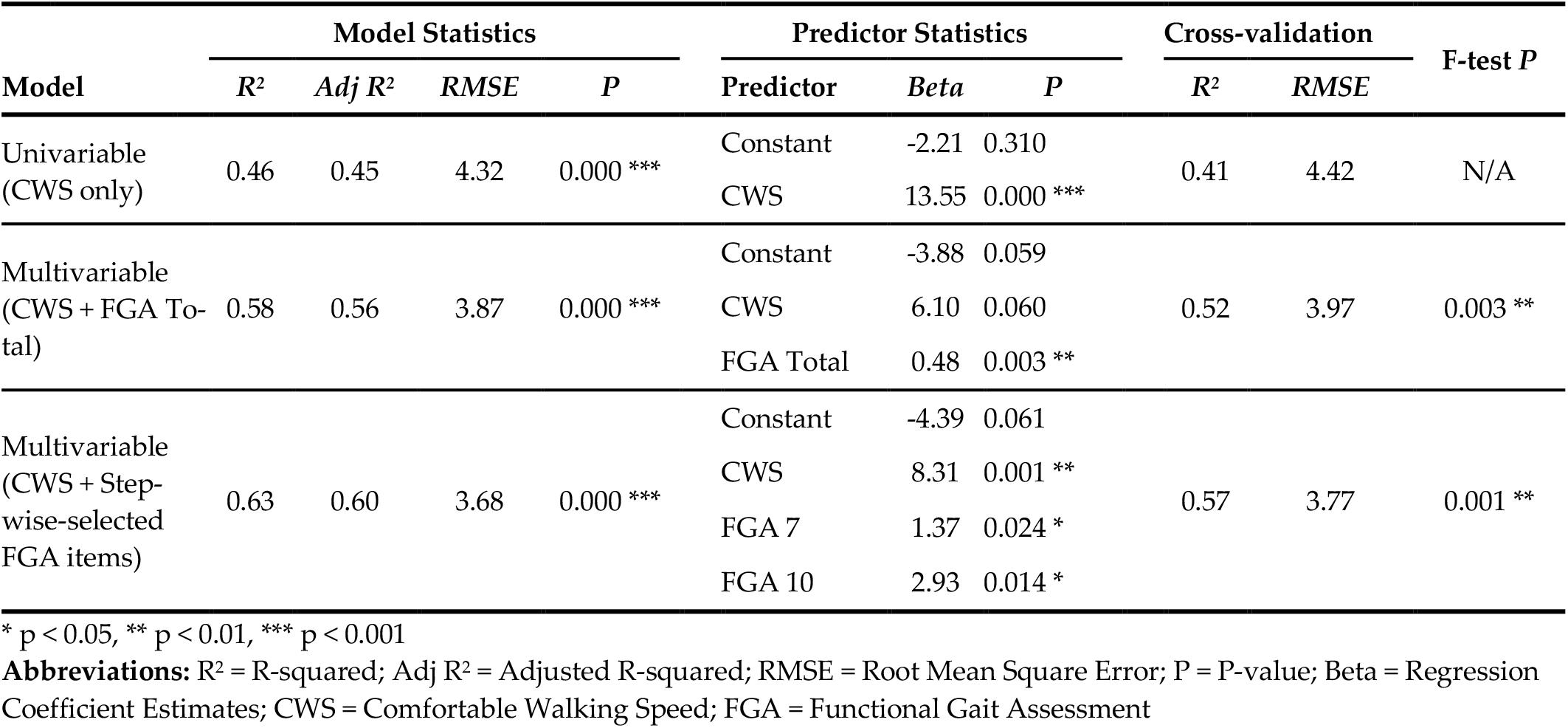
Univariable and multivariable regression analysis for the paretic propulsion peak.

Regression analysis modeling propulsion impulse is shown in Table 3. The univariable model with CWS alone explains 45% of the variance (Adj R² = 0.45, RMSE = 1.05 %BW, p < 0.001), with CWS being a significant predictor (β = 3.33, p < 0.001). The addition of the FGA Total slightly improves model fit (Adj R² = 0.49, RMSE = 1.01 %BW, p < 0.001), although the FGA Total is not statistically significant (p = 0.071) and the improvement over the CWS-only model is not significant (F(1, 37) = 3.45, p = 0.071). Multivariable model 2, selected via stepwise regression, includes CWS and FGA 10 as predictors, explaining 52% of the variance (Adj R² = 0.52, RMSE = 0.98 %BW, p < 0.001). This model significantly outperforms the CWS-only model (F(1, 37) = 6.66, p < 0.05), with all predictors reaching statistical significance (p < 0.05), and cross-validation supported improved generalizability (CV R² = 0.50, CV RMSE = 0.99 %BW).

**Table 3.** Univariable and multivariable regression analysis for paretic propulsion impulse.

| Model | Model Statistics |  |  |  | Predictor Statistics |  |  | Cross-validation |  | F-test *P* |
| --- | --- | --- | --- | --- | --- | --- | --- | --- | --- | --- |
|  | *R*<sup>2</sup> | Adj *R*<sup>2</sup> | RMSE | *P* | Predictor | Beta | *P* | *R*<sup>2</sup> | RMSE |  |
| Univariable<br>(CWS-only) | 0.47 | 0.45 | 1.05 | 0.000 *** | Constant | -0.38 | 0.468 | 0.41 | 1.08 | N/A |
|  |  |  |  |  | CWS | 3.33 | 0.000 *** |  |  |  |
| Multivariable<br>(CWS + FGA Total) | 0.51 | 0.49 | 1.01 | 0.000 *** | Constant | -0.63 | 0.232 | 0.44 | 1.05 | 0.071 |
|  |  |  |  |  | CWS | 2.20 | 0.011 * |  |  |  |
|  |  |  |  |  | FGA Total | 0.07 | 0.071 |  |  |  |
| Multivariable<br>(CWS + Stepwise-selected<br>FGA items) | 0.55 | 0.52 | 0.98 | 0.000 *** | Constant | -1.17 | 0.048 * | 0.50 | 0.99 | 0.014 * |
|  |  |  |  |  | CWS | 2.57 | 0.000 *** |  |  |  |
|  |  |  |  |  | FGA 10 | 0.75 | 0.014 * |  |  |  |

\* $p < 0.05$ , \*\* $p < 0.01$ , \*\*\* $p < 0.001$
**Abbreviations:** $R^2$ = R-squared; Adj $R^2$ = Adjusted R-squared; RMSE = Root Mean Square Error; $P$ = P-value; Beta = Regression Coefficient Estimates; CWS = Comfortable Walking Speed; FGA = Functional Gait Assessment

In contrast with regression models of paretic propulsion peak and propulsion impulse, regression models for propulsion impulse symmetry show lower predictive performance (see Table 4). The univariable model with CWS explains 20% of the variance (Adj R² = 0.20, RMSE = 0.14, p = 0.002), with a significant positive association (β = 0.26, p < 0.01). Incorporating the FGA Total does not improve model performance (Adj R² = 0.20, RMSE = 0.14), and neither predictor reaches statistical significance in the multivariable model. The improvement over the CWS-only model is also not significant (F(1, 37) = 0.91, p = 0.345). Stepwise regression does not select additional FGA items, and the resulting model is identical to the univariable model. Cross-validation results are consistent across models (CV R² = 0.17, CV RMSE = 0.14-0.15). Notably, FGA 6 is excluded from all stepwise regression analyses due to a high VIF value (VIF = 5.30).

**Table 4.** Univariable and multivariable regression analysis for propulsion impulse symmetry.

| Model | Model Statistics | | | | Predictor Statistics | | | Cross-validation | | F-test $P$ |
| --- | --- | --- | --- | --- | --- | --- | --- | --- | --- | --- |
| | $R^2$ | Adj $R^2$ | RMSE | $P$ | Predictor | Beta | $P$ | $R^2$ | RMSE | |
| Univariable<br>(CWS-only) | 0.22 | 0.20 | 0.14 | 0.002 ** | Constant | 0.12 | 0.101 | 0.17 | 0.14 | N/A |
|  |  |  |  |  | CWS | 0.26 | 0.002 * |  |  |  |
| Multivariable<br>(CWS + FGA Total) | 0.24 | 0.20 | 0.14 | 0.006 ** | Constant | 0.10 | 0.178 | 0.17 | 0.15 | 0.345 |
|  |  |  |  |  | CWS | 0.18 | 0.138 |  |  |  |
|  |  |  |  |  | FGA Total | 0.01 | 0.345 |  |  |  |
| Multivariable<br>(CWS + Stepwise-selected FGA items) | 0.22 | 0.20 | 0.14 | 0.002 ** | Constant | 0.12 | 0.101 | 0.17 | 0.14 | N/A |
|  |  |  |  |  | CWS | 0.26 | 0.002 * |  |  |  |

\* $p < 0.05$ , \*\* $p < 0.01$ , \*\*\* $p < 0.001$
**Abbreviations:** $R^2$ = R-squared; Adj $R^2$ = Adjusted R-squared; RMSE = Root Mean Square Error; $P$ = P-value; Beta = Regression Coefficient Estimates; CWS = Comfortable Walking Speed; FGA = Functional Gait Assessment

Using cutoff values of 7.02%BW and 10.86%BW, derived from tertiles of force-plate-derived paretic propulsion peak, participants may be divided into three groups: severe (< 7.02 %BW), moderate (> 7.02 %BW and < 10.86 %BW), and mild (> 10.86 %BW) groups, as shown in Figure 2a. A one-way ANOVA reveals significant differences in paretic propulsion peak across the three groups (F(2,37) = 65.60, p < 0.001). Post-hoc pairwise comparisons with Bonferroni correction confirm that all group differences are statistically significant (see Supplementary Table A1). Similarly, another one-way ANOVA finds significant differences in paretic propulsion impulse among the same groups (F(2,37) = 67.90, p < 0.001), with post-hoc Bonferroni-corrected pairwise comparisons confirming all group differences are statistically significant (see Supplementary Figure A2 and Table A2).

**Figure 2.**
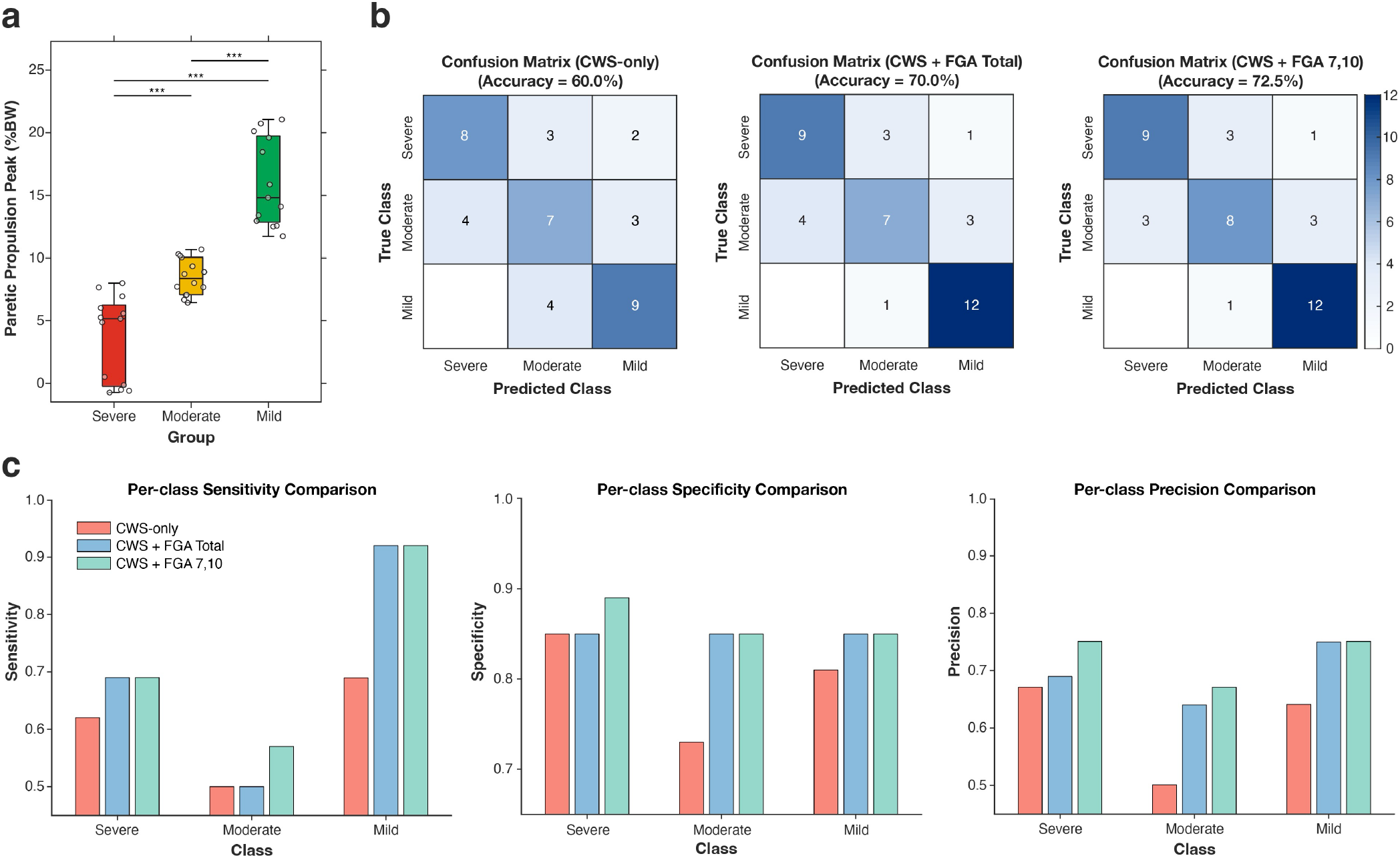
Classification of participants based on paretic propulsion peak and model performance in predicting the propulsion severity group. **(a)** Box plots illustrating group differences in paretic propulsion peak. *** indicates p < 0.001 for all pairwise comparisons; **(b)** Confusion matrices showing classification accuracy for three models predicting propulsion severity group using estimated paretic propulsion peak; **(c)** Bar plots summarizing per-class sensitivity, specificity, and precision for each model. Higher values indicated better performance across all metrics.

Confusion matrices in Figure 2b summarize the classification performance of three models in predicting the propulsion severity group using the estimated paretic propulsion peak. The model using CWS achieves an overall accuracy of 60.0%. Adding the FGA Total improves accuracy to 70.0%. The model including CWS with two stepwise-selected FGA items (FGA 7 and FGA 10) yields the highest accuracy (72.5%). Detailed per-class classification metrics are shown in Figure 2c. For the severe class, sensitivity is 0.62 in the CWS-only model and 0.69 in both the CWS + FGA Total and CWS + stepwise-selected FGA items models; specificity is 0.85, 0.85, and 0.89 across the three models; precision increases from 0.67 (CWS-only) to 0.69 and 0.75, respectively. For the moderate class, sensitivity is 0.50 in both the CWS-only and CWS + FGA Total models, and 0.57 in the CWS + stepwise-selected FGA items models; specificity improves from 0.73 to 0.85 in both FGA models; precision was 0.50, 0.64, and 0.67. For the mild class, sensitivity is 0.69 in the CWS-only model and 0.92 in both FGA models; specificity is 0.81 in the CWS-only model and 0.85 in both FGA models; and precision increases from 0.64 to 0.75 in both FGA models.

Estimation equations derived from the multivariable models were as follows, with severity groups determined as described above using cutoffs of 7.02%BW and 10.86%BW:

1. Paretic propulsion peak = -3.8817 + 6.0963 * CWS + 0.4837 * FGA Total;
2. Paretic propulsion peak = -4.3901 + 8.3050 * CWS + 1.3739 * FGA 7 + 2.9257 * FGA 10.

## 4. Discussion

Precise identification and classification of propulsion impairment is of high clinical significance; not only are propulsion deficits associated with the stability and efficiency of walking, but propulsion is a modifiable biomechanical gait subtask that, when improved, leads to meaningful increases in walking speed and functional mobility [14, 15, 16, 38].

A key observation in this study is that gait propulsion and walking speed are only moderately correlated in people post-stroke. This finding underlies why some may walk with slow gait speeds despite relatively minimal propulsion impairments, while individuals with substantial propulsion impairment may achieve a relatively fast gait speed through compensatory biomechanical strategies. This finding illustrates the limitation of using walking speed alone when classifying propulsion impairments after stroke.

Our regression analyses demonstrate that combining walking speed with clinically-derived measurements of dynamic walking ability from the FGA—captured through either the total FGA score or select FGA items—is a clinically-feasible means of characterizing propulsion impairment after stroke. Linear regression models derived from stepwise regression using CWS and FGA items # 7 (“gait with narrow base of support”) and # 10 (“steps”) offered the best quantitative prediction of paretic propulsion peak, improving classification accuracy by 12.5% compared to CWS alone. Of note, this improvement was driven primarily by enhanced differentiation among individuals with mild to moderate impairment, for whom propulsion deficits are often visually subtle. Thus, co-assessment of the FGA and walking speed extends the diagnostic precision of traditional clinic-based gait assessments and supports the broader goal of developing function-centered, biomechanical classifications of post-stroke walking ability. Interestingly, combining FGA metrics with CWS did not substantially enhance characterization of either paretic propulsion impulse or propulsion impulse symmetry. One explanation for this is that, similar to CWS, scoring the FGA is based on global performance rather than specific kinetic subtasks, which appears to limit its sensitivity to some propulsion parameters.

Other classification approaches have grouped individuals according to self-selected walking speed [14], walking activity [32], and balance self-efficacy [32]; all of these approaches consider functional mobility without regard for gait biomechanics [14, 32]. Biomechanics-informed characterization is required to inform treatment-based classifications required to guide physical rehabilitation and to distinguish between gait recovery versus compensation [15]. Classification of paretic propulsion peak using the FGA involves a function-centered examination approach that highlights biomechanical deficits associated with a variety of gait-related tasks. FGA scores have shown excellent intra-rater and interrater reliability during assessment of individuals poststroke [29]. Good to excellent concurrent validity of the FGA has been established with other measures such as the Functional Ambulation Category, Berg Balance Scale, Rivermead Mobility Index, Barthel Index, and Dynamic Gait Index [29-32]. The FGA has been shown to predict the physical activity of people poststroke and has been shown to correlate with kinematic parameters of gait [3,32]. The current findings demonstrate that the FGA—an already standardized, reliable clinical tool—can be repurposed to identify specific biomechanical deficits when analyzed through a function-centered lens. In clinical practice, this dual use preserves feasibility while enabling more precise, mechanism-informed treatment selection. Furthermore, focused subsets of the FGA may efficiently capture key biomechanical deficits when time or testing burden limit full administration. With this function-centered biomechanical classification approach, essential biomechanical impairments contributing to gait dysfunction can be identified and targeted to optimize rehabilitation and contribute to improved ambulation quality and capacity. After biomechanical capacity for gait is restored, it is possible that psychosocial and cognitive profiles that affect gait participation in community environments may be more meaningfully altered [32].

This study has several limitations. Classification thresholds were defined using sample-based tertiles of paretic propulsion peak, which may limit generalizability to populations with different propulsion distributions. In addition, this analysis focused on propulsion, while other biomechanical subtasks, such as foot clearance, single-limb support, and gait adaptability, undoubtedly contribute to overall walking function. Incorporating these elements into a unified classification system could yield a more comprehensive framework for clinical decision-making. Future studies may build on our work by examining foot clearance deficits at multiple lower-extremity joints [24]. Finally, external validation in larger and more diverse post-stroke cohorts will be essential to confirm generalizability and refine clinical thresholds.

## Conclusions

This work advances a clinically feasible and biomechanically grounded framework for classifying post-stroke gait propulsion. By demonstrating that established clinical assessments can be used to classify patients based on salient, quantitatively-derived kinetic measurements, it lays the foundation for scalable, mechanism-driven rehabilitation strategies that target the biomechanical sources of walking dysfunction.

## Author Contributions

Conceptualization, L.N.A.; Methodology, L.N.A., J.P., J.F.; Software, L.N.A., R.W., J.F.; Validation, L.N.A., J.P., J.F., R.W.; Formal Analysis, J.F., R.W., J.P.; Investigation, J.F., L.N.A.; Resources, L.N.A.; Data Curation, L.N.A, J.F., J.P., R.W.; Writing – Original Draft Preparation, J.P., J.F.; Writing – Review & Editing, J.F., J.P., L.N.A., R.W.; Visualization, J.F.; Supervision, J.P., L.N.A., J.F.; Project Administration, L.N.A., J.P., J.F.; Funding Acquisition, L.N.A.

## Institutional Review Board Statement

The study was conducted according to the guidelines of the Declaration of Helsinki and approved by the Boston University (Charles River Campus) Institutional Review Board (protocol number: #4440).

## Informed Consent Statement

Informed consent was obtained from all subjects involved in the study.

### Data Availability Statement

The data presented in this study are available on request from the corresponding author due to privacy.

## Acknowledgments

The authors acknowledge the members of the Neuromotor Recovery Laboratory for their assistance with data collection.

## Appendices

**Figure A1.**
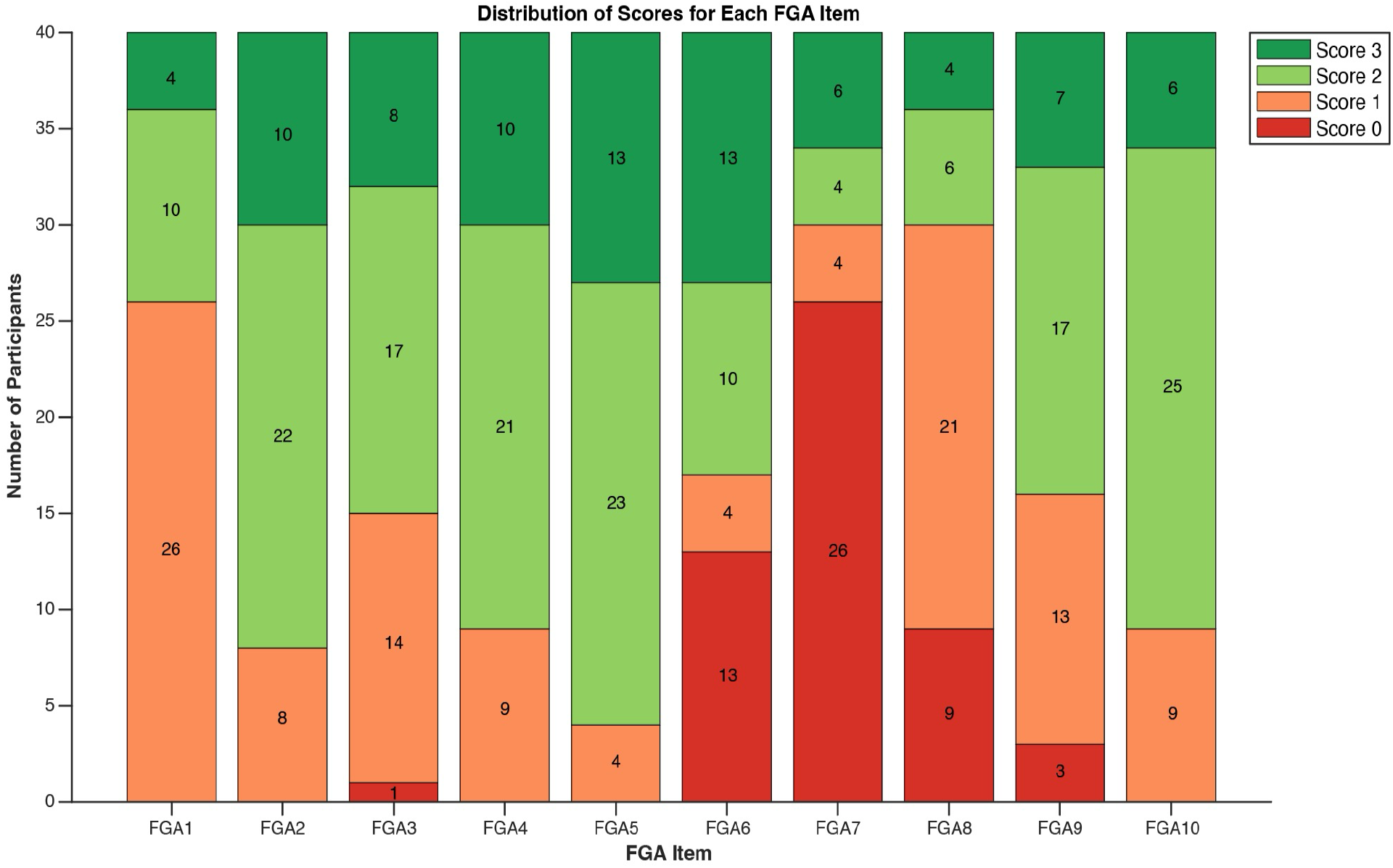
Distribution of FGA item scores among participants. Stacked bar plots show the frequency (number of participants, y-axis) for each score category (0 to 3) across the 10 FGA items (x-axis). Higher scores (green) represent better performance, while lower scores (red) indicate greater impairment.

**Table A1.** One-way ANOVA and post-hoc comparisons for paretic propulsion peak across severity groups.

| One-way ANOVA |  |  |  |  |  |
| --- | --- | --- | --- | --- | --- |
| Source | SS | DF | MS | F | P |
| Groups | 1026.00 | 2 | 513.02 | 65.60 | 0.000 *** |
| Error | 289.34 | 37 | 7.82 | - | - |
| Total | 1315.40 | 39 | - | - | - |
| Post-hoc Pairwise Comparisons |  |  |  |  |  |
| Group1 | Group2 | Mean Difference | P |  |  |
| Severe | Moderate | -5.17 | 0.000 *** |  |  |
| Severe | Mild | -12.50 | 0.000 *** |  |  |
| Moderate | Mild | -7.33 | 0.000 *** |  |  |

\* p < 0.05, \*\* p < 0.01, \*\*\* p < 0.001
**Abbreviations:** SS = Sum of Squares; DF = Degrees of Freedom; MS = Mean Square; F = F-statistic; P = P-value

**Figure A2.**
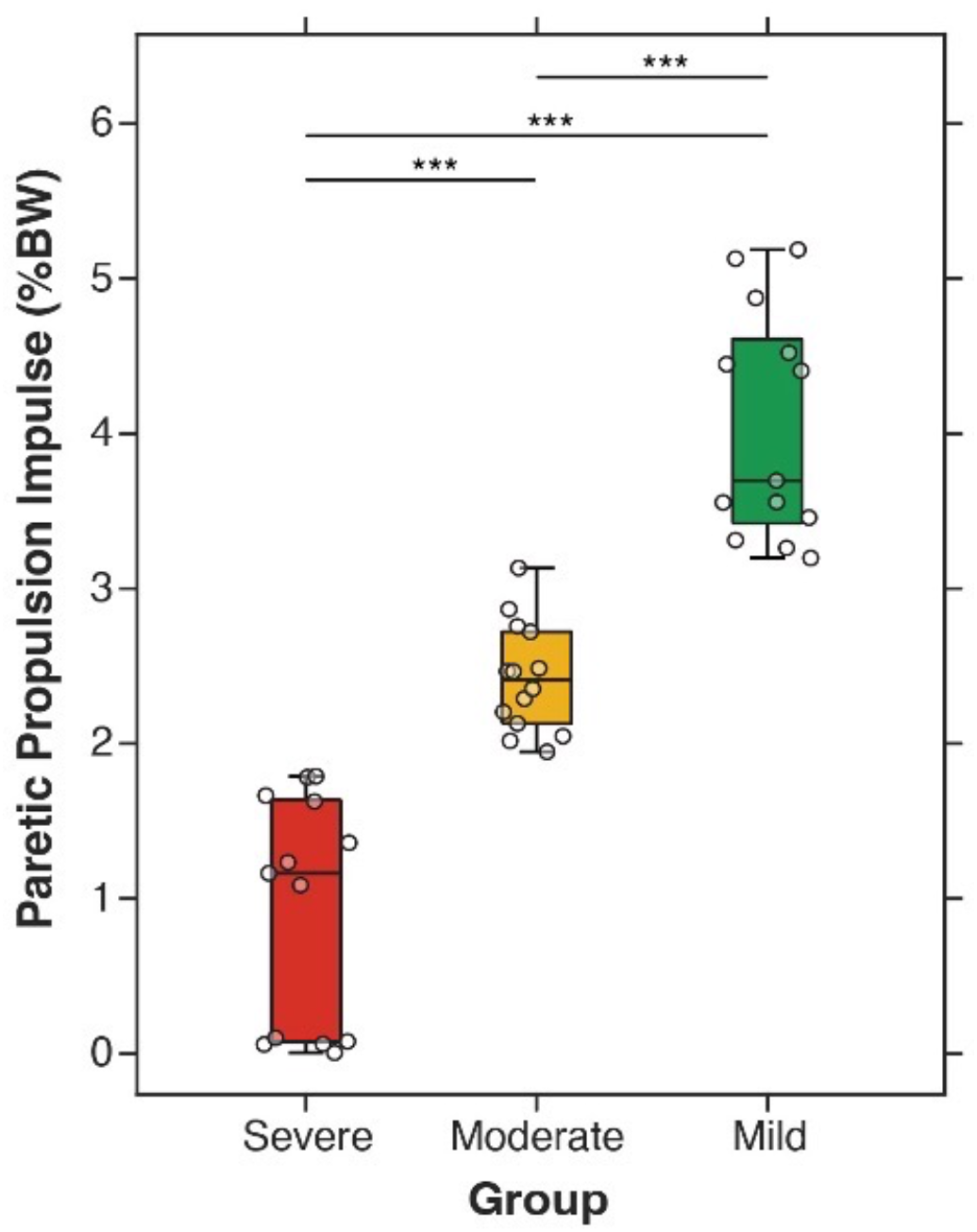
Box and whisker plots illustrating group differences in paretic propulsion impulse. *** indicates p < 0.001 for all pairwise comparisons. Three groups were categorized based on tertiles of force-plate-derived paretic propulsion peak.

**Table A2.** One-way ANOVA and post-hoc comparisons for paretic propulsion impulse across severity groups.

| One-way ANOVA |  |  |  |  |  |
| --- | --- | --- | --- | --- | --- |
| Source | SS | DF | MS | F | P |
| Groups | 61.40 | 2 | 30.70 | 67.90 | 0.000 *** |
| Error | 16.73 | 37 | 0.45 | - | - |
| Total | 78.13 | 39 | - | - | - |
| Post-hoc Pairwise Comparisons |  |  |  |  |  |
| Group1 | Group2 | Mean Difference | P |  |  |
| Severe | Moderate | -1.39 | 0.000 *** |  |  |
| Severe | Mild | -3.07 | 0.000 *** |  |  |
| Moderate | Mild | -1.68 | 0.000 *** |  |  |

\* $p < 0.05$ , \*\* $p < 0.01$ , \*\*\* $p < 0.001$
**Abbreviations:** SS = Sum of Squares; DF = Degrees of Freedom; MS = Mean Square; F = F-statistic; P = P-value

